# Inequalities in COVID19 mortality related to ethnicity and socioeconomic deprivation

**DOI:** 10.1101/2020.04.25.20079491

**Authors:** Tanith C. Rose, Kate Mason, Andy Pennington, Philip McHale, Iain Buchan, David C. Taylor-Robinson, Ben Barr

**Affiliations:** Department of Public Health and Policy, University of Liverpool, L69 3GL

## Abstract

**Background:** Initial reports suggest that ethnic minorities may be experiencing more severe coronavirus disease 2019 (COVID19) outcomes. We therefore assessed the association between ethnic composition, income deprivation and COVID19 mortality rates in England.

**Methods:** We performed a cross-sectional ecological analysis across England’s upper-tier local authorities. We assessed the association between the proportion of the population from Black, Asian and Minority Ethnic (BAME) backgrounds, income deprivation and COVID19 mortality rates using multivariable negative binomial regression, adjusting for population density, proportion of the population aged 50–79 and 80+ years, and the duration of the epidemic in each area.

**Findings:** Local authorities with a greater proportion of residents from ethnic minority backgrounds had statistically significantly higher COVID19 mortality rates, as did local authorities with a greater proportion of residents experiencing deprivation relating to low income. After adjusting for income deprivation and other covariates, each percentage point increase in the proportion of the population from BAME backgrounds was associated with a 1% increase in the COVID19 mortality rate [IRR=1.01, 95%CI 1.01–1.02]. Each percentage point increase in the proportion of the population experiencing income deprivation was associated with a 2% increase in the COVID19 mortality rate [IRR=1.02, 95%CI 1.01–1.04].

**Interpretation:** This study provides evidence that both income deprivation and ethnicity are associated with greater COVID19 mortality. To reduce these inequalities, Government needs to target effective control and recovery measures at these disadvantaged communities, proportionate to their greater needs and vulnerabilities, during and following the pandemic.

**Funding:** National Institute of Health Research; Medical Research Council

## Introduction

To date, the coronavirus disease 2019 (COVID19) pandemic has claimed thousands of lives in the UK. The admission of Boris Johnson, the UK Prime Minister, to hospital prompted some to portray the disease as a great equaliser, which can affect anyone, even the most privileged. But are all communities equally affected? There is growing evidence that the adverse consequences of the pandemic are falling disproportionately on more disadvantaged groups, particularly those from ethnic minorities.

Evidence emerging from the United States shows that COVID19 mortality is disproportionately high amongst African Americans and other ethnic minority communities. In Chicago, around 70% of deaths have involved people of Black origin despite them making up only 30% of the population, and similar figures have been observed in several other states.^1^ ^2^ Estimates suggest that American counties where Black residents are in the majority have almost six times the rate of death due to COVID19 compared to counties with predominantly White residents.^2^ In the UK, COVID19 deaths have only recently been reported by ethnic group, showing that 19% of deaths occurring in hospital have involved individuals from ethnic minority backgrounds, even though these groups make up only 14% of the population.^3^ Similarly Black, Asian and Minority Ethnic (BAME) groups accounted for 35% of all COVID19 patients in critical care units in England.^4^

BAME groups may be at greater risk of infection, severe disease and poor outcomes for multiple reasons. These include socioeconomic conditions that increase risk of transmission and vulnerability^5 6^ (e.g. overcrowded housing, employment in essential occupations, poverty and reliance on public transport), unequal access to effective healthcare and higher rates of comorbidities, such as diabetes, hypertension, and cardiovascular diseases.^7^ These comorbidities have all been associated with COVID19 mortality^8 9^ and are also more common in BAME groups.^10,11^

The extent to which the higher COVID19 mortality in BAME groups is primarily due to socioeconomic inequalities is not fully understood and there has been little or no research investigating whether poverty itself is an independent risk factor for COVID19 mortality. The same risks outlined above, e.g. household overcrowding, reliance on public transport, working in essential occupations and increased comorbidities, are also associated with living in poverty, as well as ethnicity. It is challenging to disentangle the potential impacts of socioeconomic factors and ethnicity on adverse health outcomes and there has been much debate, in the UK, about whether ethnic inequalities in health in general are primarily explained by socioeconomic inequalities.^12^ ^13^ Ethnic minority groups in the UK are more likely to live in socioeconomically deprived neighbourhoods compared to the White British population, and are more likely to experience poverty and unemployment.^14 15^

It is important to understand the relationship between ethnicity, poverty, and mortality due to COVID19 in order that resources and control measures can be better targeted at the communities most at risk – now and in the aftermath of the pandemic. We do not know, however, the extent to which these factors explain differences in risk between communities in England. To address this gap, we investigated the association between ethnic composition, income deprivation and COVID19 mortality rates across areas of England, whilst controlling for population density, the age composition of the population and the duration of the epidemic in each area.

## Methods

### Study design and setting

We performed a cross-sectional ecological analysis across upper-tier local authorities in England. Upper-tier local authorities are administrative municipality areas, each containing an average population of around 370,000 people. We merged two local authorities (Dorset with Bournemouth, Christchurch and Poole) which were subject to administrative changes between 2018 and 2019, to enable linkage of datasets produced at different time points. We also merged Isles of Scilly with Cornwall due to the small population size of the former and excluded two local authorities (City of London and Rutland) due to the small number of confirmed cases in these areas, leaving 147 upper-tier local authorities for analysis.

### Data sources and measures

To derive our measure of COVID19 mortality, we used data provided by NHS England on deaths of patients who died in hospitals who tested positive for COVID19 at time of death, recorded up until the 22^nd^ April 2020.^3^ Data are provided with deaths aggregated by NHS Trust (individual NHS hospitals or small groups of hospitals in England serving a geographical area).

In order to map these hospital level data to local authority we used 2018/19 data on all-cause emergency hospital admissions aggregated by NHS Trust and local authority of patient residence, to calculate the proportion of each NHS Trust’s admissions that have historically come from each local authority area. We then applied this proportion (weight) to the mortality data assuming that deaths from each NHS Trust were distributed between local authorities based on the historical share of admissions from that local authority.

We used these hospital data as they are the most up-to-date statistics available on COVID19 mortality in England. There is a long time-lag in the reporting of more comprehensive data from the Office for National Statistics (ONS), which also includes deaths outside hospital. As a robustness test however, we replicated the analysis using the most recent ONS mortality figures up to 10^th^ April 2020.

We measured the proportion of the population from a BAME group in each local authority reporting their ethnic group as Black, Asian, Mixed or Other, in the 2011 Census. Our measure of poverty, was the income deprivation domain from the Index of Multiple Deprivation (IMD) 2019 score, provided by the Ministry of Housing, Communities & Local Government. The IMD score is an overall measure of socioeconomic deprivation experienced by people living in a local authority. The income deprivation score measures the percentage of the population experiencing deprivation relating to low income, based on a non-overlapping count of people receiving welfare benefits for low-income. It includes those who are out-of-work and those who are in work but have low earnings.^16^

Population estimate data provided by the ONS were used to calculate population densities and the percentage of the population aged 50–79 years and 80+ years per local authority. Finally, to control for differences in the duration of the epidemic in each local authority, we used Public Health England data to calculate the number of days from when each local authority had at least 10 laboratory confirmed COVID19 cases to the 22^nd^ April.

### Statistical Analysis

The statistical analysis proceeded in two steps. First we performed descriptive analyses to explore the correlation between each local authority’s COVID19 mortality rate and (1) the proportions of people from BAME backgrounds and (2) income deprivation in each local authority. Second, we used multivariable regression to investigate how these associations changed when controlling for population density, proportion of the population aged 50–79 and 80+ years, and the duration of the epidemic in each area.

We used negative binomial regression models to account for overdispersion of the data (mortality rate variance likely to exceed the mean, breaking Poisson assumptions). Numbers of deaths were modelled using the log of the population size as an ‘offset’ variable, effectively modelling the log of the rate (see Supplementary file for full details of the statistical model). We conducted several sensitivity analyses, repeating our analysis using linear regression to model the mortality rate, using ONS mortality figures up to 10^th^ April 2020 and excluding local authorities in London. We also investigated the association between the proportion of the population from Black ethnic backgrounds and COVID19 mortality. Analyses were conducted using R (version 3.6.0) and Stata (version 13).

## Role of the funding source

The funders of the study had no role in study design, data collection, data analysis, data interpretation, or the writing of the report. The authors had full access to all of the data in the study and the final responsibility to submit for publication.

## Results

Figure 1 illustrates the crude linear association between the proportion of people from BAME backgrounds and the COVID19 mortality rate in each local authority. Local authorities, where a larger proportion of the population were from BAME backgrounds, tended to have higher COVID19 mortality rates. Areas that were more income deprived also tended to have higher COVID19 mortality rates although this association was weaker (Figure 2).

**Figure 1:**
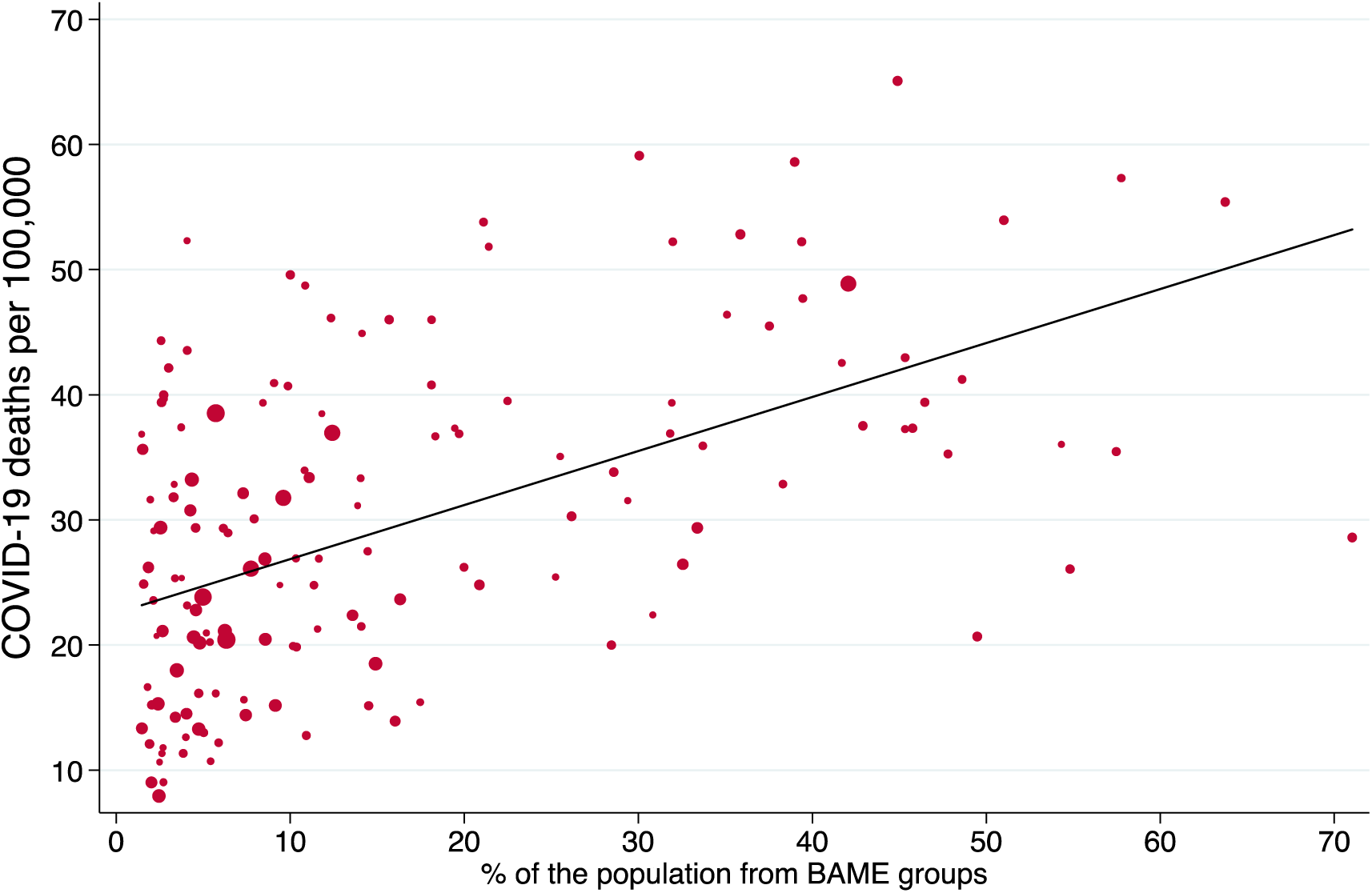
Linear association between the percentage of people from BAME backgrounds and the COVID19 mortality rate for local authorities in England. The size of each data point is proportional to the local authority population.

**Figure 2:**
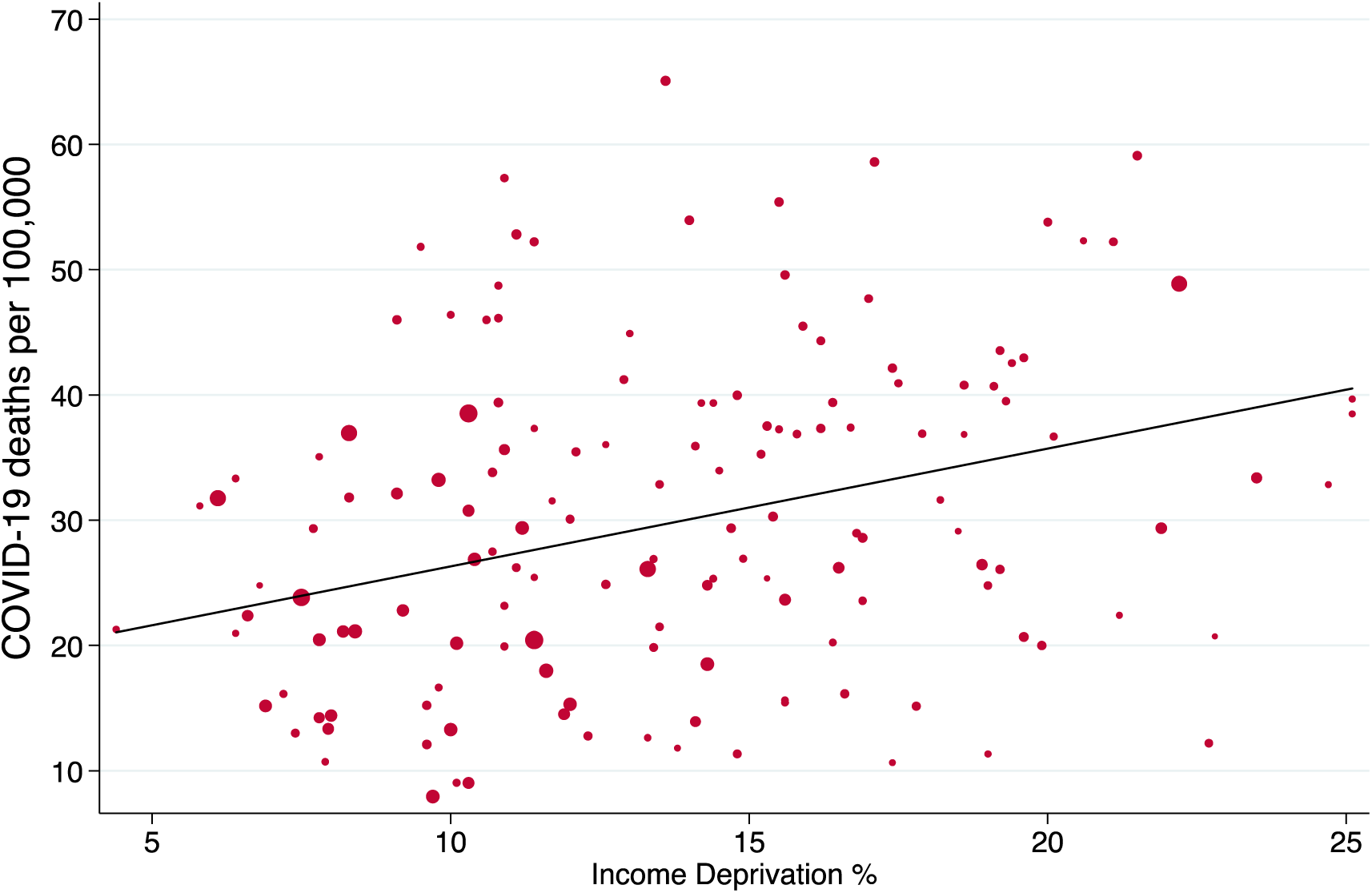
Linear association between the income deprivation score and the COVID19 mortality rate for local authorities in England. The size of each data point is proportional to the local authority population.

The multivariable regression analyses showed that the associations in Figures 1 and 2 remained after adjusting for population density, the duration of the epidemic and the proportion of older residents (Table 1). Both ethnicity and income deprivation were independently associated with COVID19 mortality. Results from the fully adjusted model (Model 3) showed that each 1 percentage point increase in the proportion of the population from BAME backgrounds was associated with a 1% increase in the COVID19 mortality rate [IRR=1.01, 95%CI 1.01 to 1.02]. Furthermore, each 1 percentage point increase in the proportion of the population experiencing deprivation relating to low income was associated with a 2% increase in the COVID19 mortality rate [IRR=1.02, 95%CI 1.01 to 1.04]. Results from sensitivity analyses all indicated similar findings - using ONS mortality figures, linear regression and models excluding London (Supplementary file). The adjusted incident rate ratio of mortality per percentage point increase in the proportion of the population from Black ethnic backgrounds was 1.026 (95%CI 1.01 to 1.04) (Supplementary file).

**Table 1:** Multivariable negative binomial regression models for COVID19 mortality rate for local authorities in England.

|  | Model 1 |  |  | Model 2 |  |  | Model 3 |  |  |
| --- | --- | --- | --- | --- | --- | --- | --- | --- | --- |
|  | IRR | [95%<br>Conf | Interval] | IRR | [95%<br>Conf | Interval] | IRR | [95%<br>Conf | Interval] |
| Duration (days) | 1.000 | 0.985 | 1.014 | 1.018** | 1.001 | 1.036 | 1.010 | 0.994 | 1.027 |
| Aged 50-79 (%) | 1.031* | 0.995 | 1.069 | 1.002 | 0.969 | 1.037 | 1.030 | 0.994 | 1.067 |
| Aged 80+ (%) | 0.917 | 0.800 | 1.051 | 0.936 | 0.813 | 1.077 | 0.937 | 0.820 | 1.071 |
| Population density<br>(10,000 pop/km<sup>2</sup>) | 1.157 | 0.842 | 1.591 | 1.183 | 0.846 | 1.655 | 1.068 | 0.779 | 1.465 |
| BAME (%) | 1.014*** | 1.007 | 1.022 | - | - | - | 1.014*** | 1.007 | 1.021 |
| Deprivation (%) | - | - | - | 1.024*** | 1.006 | 1.042 | 1.022*** | 1.006 | 1.039 |

Models based on 147 observations (full model results are in Supplementary file)
BAME = Black, Asian and Minority Ethnic; IRR = incident rate ratio
\*\*\* $p < 0.01$ , \*\* $p < 0.05$ , \* $p < 0.1$

## Discussion

We explored the relationships between ethnicity, deprivation and COVID19 mortality at upper-tier local authority level in England, and revealed stark ethnic and socioeconomic inequalities in mortality rates. Local authorities with a greater proportion of residents from BAME groups had higher COVID19 mortality rates, as did local authorities with a greater proportion of residents experiencing deprivation from low income. These associations were independent of each other, and remained after controlling for population density, the duration of the epidemic and the proportion of older residents. These findings corroborate previous evidence that ethnic minority groups are at greater risk of COVID19 mortality. They indicate, at least in aggregated geographies, that these ethnic inequalities do not appear to be fully explained by socioeconomic differences. For the first time in the UK, we demonstrate that poverty appears to be an independent risk factor for COVID19 mortality.

These findings could relate to differences in exposure, differences in susceptibility or differences in severe consequences from infection to COVID19, by ethnicity and income deprivation.^17^ Differences in exposure could relate to factors such as overcrowded housing. Statistics from the English Housing Survey suggest that 30% of Bangladeshi households experience overcrowding, as do 16% of Pakistani and 12% of Black households, compared to 2% of White British households.^5^ The prevalence of overcrowding also increases with socioeconomic disadvantage for both ethnic minority and White British households.^5^ Overcrowding, which may be related to multi-generational living arrangements, is likely to pose significant challenges for households trying to control the spread of infection. Employment conditions may also increase risk of exposure for disadvantaged populations as they have less employment protection and may be less likely to be able to work from home and follow social distancing guidelines. The self-employed and those who earn less than £118 per week are not entitled to receive Statutory Sick Pay in the UK.^18^ Taking sickness absence may therefore incur loss of earnings and financial penalties for those who can ill afford it.

Part of the increased susceptibility could relate to socially patterned factors that impair immune response, including increased levels of chronic stress, smoking, obesity and nutritional deficiencies.^19 20^ ^21^ For example, studies have found that individuals of lower socioeconomic status compared to high are more routinely exposed to psychosocial stressors in their living and working environments, contributing to a persistent level of background stress.^22^ Experiences of racial harassment and discrimination may also be important sources of psychological distress for ethnic minorities.^12^ Chronic stress has been shown to have negative effects on immune system functioning, suppressing the body’s ability to initiate an efficient immune response to infection.^19^

Differences in the severity and consequences of disease could relate to patterns of pre-existing chronic conditions which are more common amongst ethnic minorities and disadvantaged communities. Studies have reported elevated risk of mortality and other severe COVID19 outcomes associated with hypertension, coronary heart disease and diabetes,^8^ ^9^ ^23^ which are more prevalent amongst BAME groups and people living in poverty.^10^ ^11^ ^24^ Differences in how these conditions are managed might also play a role, for example ACE-inhibitors are not recommended for the treatment of patients from Black ethnic backgrounds with hypertension, but emerging evidence suggests ACE-inhibitors may be protective against COVID19 mortality.^25^

Variation in healthcare seeking behaviours and access to healthcare amongst more disadvantaged groups could also contribute to differences in the consequences of COVID19. Some studies have found that ethnic minorities are more likely to use general practice services compared to the White British population, even after controlling for their increased levels of need.^26^ Yet surveys of patients’ experiences of healthcare in the UK have shown that ethnic minorities tend to report lower levels of satisfaction and less positive experiences of care.^27^ ^28^ Furthermore, over the last decade the UK government has implemented a series of ‘hostile environment’ policies for migrants, which included a requirement for the NHS to alert authorities to people suspected of being in the country illegally, and these may have influenced the extent to which migrants seek healthcare when in need.^29^

### Strengths and limitations

This study reveals, for the first time, ethnic and socioeconomic inequalities in COVID19 mortality rates across English upper-tier local authorities. We performed a comprehensive analysis of all COVID19 deaths of patients in English hospitals, using the most up-to-date data readily available, therefore our results are likely to be broadly generalisable to the English population, although caveats regarding the underreporting of deaths within official figures should be considered. Additionally, a number of robustness tests confirmed our results, using alternative models, less up-to-date but more comprehensive ONS death data, and models excluding London. Our research highlights an important emerging issue, and it is intended that the findings will prompt further investigation.

Our results are nonetheless preliminary, and there are several limitations that should be considered when interpreting the results. In terms of study design, methodological limitations of ecological studies can include ecological bias whereby associations present at the group-level are not apparent at the individual-level, possibly due to unmeasured confounding or measurement error.^30^ Data were aggregated to relatively large areas (upper tier local authorities containing approximately 370,000 people on average) which may have increased the likelihood of ecological bias. Nevertheless, because ecological studies are able to capture risk factors and exposures that operate at the community-level,^31^ it could be argued that ecological studies are in fact more appropriate for the study of infectious diseases compared to individual-level studies. Individual-level studies will however be required to understand the causal pathways leading to these inequalities we observe.

Analysis was performed on 147 local authorities, which provides a limited number of data points, meaning that our analysis is relatively underpowered. In our analysis, we also made the assumption that deaths from each NHS Trust were distributed between local authorities based on the historical share of hospital admissions from that local authority. Whether this affected the results is unknown, however when we repeated the analysis using ONS mortality figures based on local authority of residence we found very similar results (Supplementary file).

Adjustment for duration of the pandemic assumed a similar shaped epidemic curve, i.e. exposure per unit time in each locality, however we acknowledge that earlier affected local authorities, before lock-down, particularly in London, will have experienced steeper curves. Additionally, we have focused on the shared potential wider determinants of COVID19 mortality across people from BAME groups, however supplementary analyses revealed a stronger association between the proportion of the population from Black ethnic backgrounds and COVID19 mortality, compared to the association for all BAME groups combined. Further research is needed, using both quantitative and qualitative methods to disentangle and understand the pathways though which ethnicity and deprivation influence mortality.

## Implications for policy

Our findings have important implications for policies that aim to minimise the adverse consequences of the pandemic and reduce its impact on health inequalities. Inequalities in health in the UK were increasing in the decade before the COVID19 pandemic.^32^ Our findings suggest that the current crisis will increase these inequalities further, unless urgent action is taken. To have the greatest impact on reducing inequalities, resources need to be delivered in proportion to the degree of need – sometimes referred to as proportionate universalism.^24^ Current approaches, however, are not sufficiently taking into account the key drivers of need, that we identify – poverty and ethnicity.^33^ The centralised and universal implementation of control measures, that has been applied so far, will not address these inequalities. What is needed is an approach that is tailored to the communities most at risk, led by local government in partnership with these communities and informed by granular intelligence to rapidly identify problems early and rapidly target responses. This is particularly important as we move into a phase of testing and tracing. To address the higher risks we have shown amongst people living in poverty and ethnic minority populations, this will need to involve organisations that are trusted in those communities. Over reliance on technical solutions such as contact tracing apps, may widen these inequalities as they are taken up preferentially by more advantaged groups. Our study indicates that the adverse effects of COVID19 are falling disproportionately on people living in poverty and people from ethnic minority groups and if action is not taken urgently to address this, we will potentially loose another decade in the fight against health inequalities.

## Data Availability

All data are publicly available

## Acknowledgments

BB and TR are supported by the National Institute for Health Research (NIHR) Applied Research Collaboration North West Coast (ARC NWC). BB, KM and DTR are supported by the NIHR School for Public Health Research. IB is supported by NIHR Senior Investigator award. This report is independent research funded by the NIHR ARC NWC. The views expressed in this publication are those of the author(s) and not necessarily those of the NIHR or the Department of Health and Social Care. PM is funded through an MRC Clinical Research Training Fellowship (MR/T00794X/1). DTR is funded by the MRC on a Clinician Scientist Fellowship (MR/P008577/1).

## Conflict of interest statement

All authors have completed the ICMJE uniform disclosure format www.icmje.org/coi_disclosure.pdf and declare: BB and TR are supported by the NIHR Applied Research Collaboration North West Coast; BB, KM and DTR are supported by the NIHR School for Public Health Research; PM and DTR are funded by the Medical Research Council; IB is supported by NIHR Senior Investigator award; no financial relationships with any organisations that might have an interest in the submitted work in the previous three years; no other relationships or activities that could appear to have influenced the submitted work.

## Authors’ contributions

BB conceived the study. BB and TR conducted the analysis. TR and BB drafted the manuscript with DTR. KM, AP and PM performed literature searches. All authors interpreted the data and revised the manuscript.

## Ethics committee approval

None required

## Notes

### Competing Interest Statement

The authors have declared no competing interest.

